# Distinct immune signatures discriminate SARS-CoV-2 vaccine combinations

**DOI:** 10.1101/2022.09.05.22279572

**Authors:** Nicolás Gonzalo Núñez, Jonas Schmid, Laura Power, Chiara Alberti, Sinduya Krishnarajah, Stefanie Kreutmair, Susanne Unger, Sebastián Blanco, Brenda Konigheim, Constanza Marín, Luisina Onofrio, Jenny Christine Kienzler, Sara da Costa Pereira, Florian Ingelfinger, InmunoCovidCba, InViV working group, Marina E. Pasinovich, Juan M Castelli, Carla Vizzotti, Maximilian Schaefer, Juan Villar-Vesga, Carla Helena Merten, Aakriti Sethi, Tobias Wertheimer, Mirjam Lutz, Danusia Vanoaica, Claudia Sotomayor, Adriana Gruppi, Christian Münz, Diego Cardozo, Gabriela Barbás, Laura Lopez, Paula Carreño, Gonzalo Castro, Elias Raboy, Sandra Gallego, Gabriel Morón, Laura Cervi, Eva V Acosta Rodriguez, Belkys A Maletto, Mariana Maccioni, Burkhard Becher

## Abstract

Several vaccines have been found effective against COVID-19, usually administered in homologous regimens, with the same vaccine used for the prime and boost doses. However, recent studies have demonstrated improved protection via heterologous mix-and-match COVID-19 vaccine combinations, and a direct comparison among these regimens is needed to identify the best employment strategies. Here, we show a single-cohort comparison of changes to the humoral and cellular immune compartments following five different COVID-19 vaccines spanning three technologies (adenoviral, mRNA and inactivated vaccines). These vaccines were administered in a combinatorial fashion, resulting in sixteen different homologous and heterologous regimens. SARS-CoV-2-targeting antibody titres were highest when the boost dose consisted of mRNA-1273, independent of the vaccine used for priming. Priming with BBIBP-CorV induced less class-switching among spike-binding memory B cells and the highest antigen-specific T cell responses in heterologous combinations. These were generally more immunogenic in terms of specific antibodies and cellular responses compared to homologous regimens. Finally, single-cell analysis of 754 samples revealed specific B and T cell signatures of the vaccination regimens, indicating distinctive differences in the immune responses. These data provide new insights on the immunological effects of COVID-19 vaccine combinations and a framework for the design of improved vaccination strategies for other pathogens and cancer.

## Main text

The management of the COVD-19 pandemic has relied heavily on the development and global deployment of vaccines that protect against SARS-CoV-2. Numerous homologous prime-boost vaccination regimens were shown to stimulate robust cellular and humoral immune responses and have been approved for clinical use^1,2^. The many different vaccines and vaccine technologies employed during this pandemic provide a unique opportunity to study their effects and dissect the immune mechanisms of protection associated with each regimen. Recent reports have shown that certain heterologous prime-boost regimens provide enhanced protection against SARS-CoV-2^3–13^ (https://www.who.int/publications/i/item/WHO-2019-nCoV-vaccines-SAGE-recommendation-heterologous-schedules), in line with preclinical data comparing heterologous vs. homologous vaccination regimens against numerous other pathogens^14–22^. Nevertheless, because the corresponding differences in antibody and cellular immune responses are not fully understood, there is still no consensus on the most appropriate regimens for existing COVID-19 vaccines. Furthermore, little is known about the immunogenicity and reactogenicity of heterologous combinations of most COVID-19 vaccines available on the market.

The primary COVID-19 vaccination regimens approved for clinical use include viral-vector based approaches such as AZD1222 (two-dose homologous prime-boost, henceforth termed AZD), developed by the University of Oxford in collaboration with AstraZeneca^23,24^; Sputnik V (prime-boost heterologous human adenoviral vectors, serotypes 26 (henceforth Sput 26) and 5 (Sput 5)) by the Gamaleya Research Institute^2,25^; and Ad5-nCoV-S (a single-dose viral vector vaccine, henceforth Ad5) by CanSino Biologics^26,27^. Also included are inactivated vaccines such as BBIBP-CorV (henceforth BBIBP), developed by Sinopharm^28^, and mRNA-based vaccines such as mRNA-1273 by Moderna^29,30^, as well as protein subunit vaccines such as NVX-CoV2373, a recombinant spike protein nanoparticle vaccine by Novavax^31^. These vaccines are being widely administered on a global scale (https://covid19.trackvaccines.org/agency/who/).

In 2021, the Argentine Ministry of Public Health began a multicentre study to systematically compare the immune responses to homologous and heterologous COVID-19 vaccination regimens in Argentina^32^. In the present study, we specifically analysed samples from one cohort from ECEHeVac (NCT04988048) located at a clinical centre in the city of Córdoba (REPIS-Cba #4371). The individuals recruited for the trial were first vaccinated with one dose of AZD, BBIBP, Sput 26, or mRNA-1273, followed by a randomized second dose of AZD, BBIBP, Sput 26, Sput 5, mRNA-1273 or Ad5, resulting in a total of 16 different homologous or heterologous prime-boost regimens. We conducted a head-to-head comparison of immune responses to COVID-19 vaccine regimens and interrogated antibody and cellular responses, which are known to act in tandem to provide protection against SARS-CoV-2^33,34^, for all 16 prime-boost regimens and correlated them with their immunogenicity.

### Study design and vaccine combinations

To compare the immunological changes resulting from different COVID-19 vaccination regimens, we enrolled subjects to be vaccinated with the regimens listed in **Fig. 1a**. Individuals were given a priming dose (Dose 1) followed by a homologous or heterologous booster dose (Dose 2) 4-12 weeks later, resulting in a total of 16 combinatorial vaccination regimens. The interval between Dose 1 and Dose 2 depended on the first vaccine administered. Blood samples were collected from each volunteer at 4-12 weeks after Dose 1 (T1), 14 ± 2 days after Dose 2 (T2), and 28 ± 1 days after Dose 2 (T3) (**Fig. 1a-b**). The T1-T2 and T2-T3 intervals were approximately two weeks for all groups (**Fig. 1b**). Serum and plasma were collected at all timepoints, and peripheral blood mononuclear cells (PBMCs) were isolated at T1 and T3. In total, 1,491 samples from 497 individuals were included in the analysis. Characteristics of the whole cohort and the vaccine groups are summarized in **Fig. 1b**. Individuals with a prior history of COVID-19 or a positive SARS-CoV-2 nucleocapsid protein IgG ELISA result at T1 were excluded. The vaccine groups were comparable in number of participants and gender distribution.

**Figure 1.**
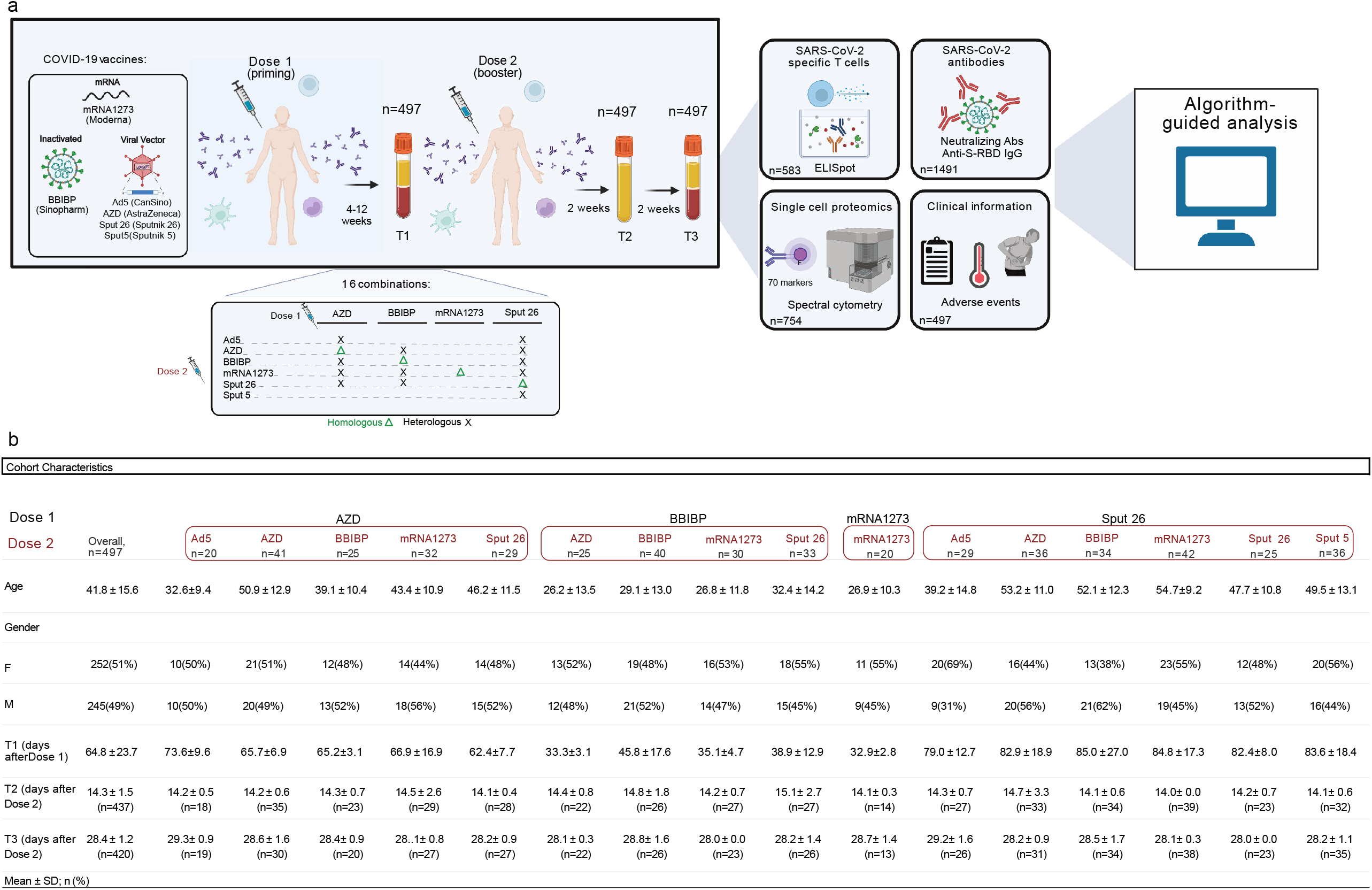
Schematic of vaccination regimens and data analysis pipeline. (a)Schematic representation of the study protocol. Participants were vaccinated with one of 16 COVID-19 vaccine combination regimens and donated blood at timepoints T1 (4-12 weeks after Dose 1), T2 (two weeks after Dose 2), and T3 (four weeks after Dose 2). PBMCs collected at T1 and T3 were used for IFNγ ELISpot assays and high-dimensional spectral flow cytometry analysis. Sera and plasma collected at T1, T2, and T3 were analysed for anti-S-RBD IgG levels and NAb titres, respectively. Participants were monitored for adverse events. The data collected were analysed using dimensionality reduction, FlowSOM-based clustering algorithms, and statistical testing. (b) Cohort characteristics. Mean number of days and standard deviation after Dose 1 or Dose 2 were calculated from a subset of the total participants, as indicated below each mean.

All study participants were monitored for up to 6 months after Dose 2 to assess reactogenicity. Pain at the site of injection was the most frequent local adverse event after Dose 1 (**Extended Data Fig. 1a**) and Dose 2 (**Extended Data Fig. 1b**). The most common systemic adverse manifestations were fever, arthromyalgia, and headache. In general, the spectrum of reported adverse events was comparable across all groups. Of note, the use of mRNA-1273 or Sput 26 as Dose 2 in heterologous regimens, and the combination of AZD/Ad5 (Dose 1/Dose 2), increased the frequency of reported cumulative adverse events compared to homologous regimens. All reactogenicity symptoms were short-lived, and there were no hospitalizations due to adverse events. Thus, all vaccine regimens were considered collectively well-tolerated.

### mRNA-1273 boost enhances humoral response

We evaluated the antibody response to the 16 prime-boost regimens by measuring participants’ serum levels of IgG specific for the receptor-binding domain (RBD) of the SARS-CoV-2 spike protein (anti-S-RBD IgG) and their SARS-CoV-2-neutralizing antibody (NAb) titres at timepoints T1, T2, and T3 for all vaccination groups (**Fig. 2a-b**). All study participants had detectable anti-S-RBD IgG levels (> 50 AU/ml) after Dose 2, and anti-S-RBD IgG levels increased in most vaccine groups at T2 and T3 compared to T1 (**Fig. 2a**). Anti-S-RBD IgG levels increased significantly from T1 to T3 in all groups except for AZD/BBIBP. Peak antibody concentrations were reached at T2, with a marginal and non-significant decrease at T3, for almost all groups (**Fig. 2a**). The NAb titres showed similar patterns, although detectable titres (≥ 1:10) were not present in all individuals (**Fig. 2b**). Both of these humoral responses were most strongly enhanced by mRNA-1273 except after priming with AZD, where boosting with Ad5 led to the greatest fold increase in NAb titres (**Extended Data Fig. 2a-b**). At T3, anti-S-RBD IgG levels and NAb titres were highest in individuals that received homologous mRNA-1273/mRNA-1273 vaccination and in all the heterologous groups that included mRNA-1273 as Dose 2 (**Fig. 2c-d**). Anti-S-RBD IgG levels were also positively correlated with NAb titres at T3 (**Extended Data Fig. 2c**). Thus, heterologous vaccine combinations generally elicited enhanced antibody responses compared to homologous combinations, except when BBIBP was administered as Dose 2 (**Fig. 2c-d**). In conclusion, using mRNA-1273 as Dose 2 induced the strongest antibody responses, regardless of the vaccine used as Dose 1.

**Figure 2.**
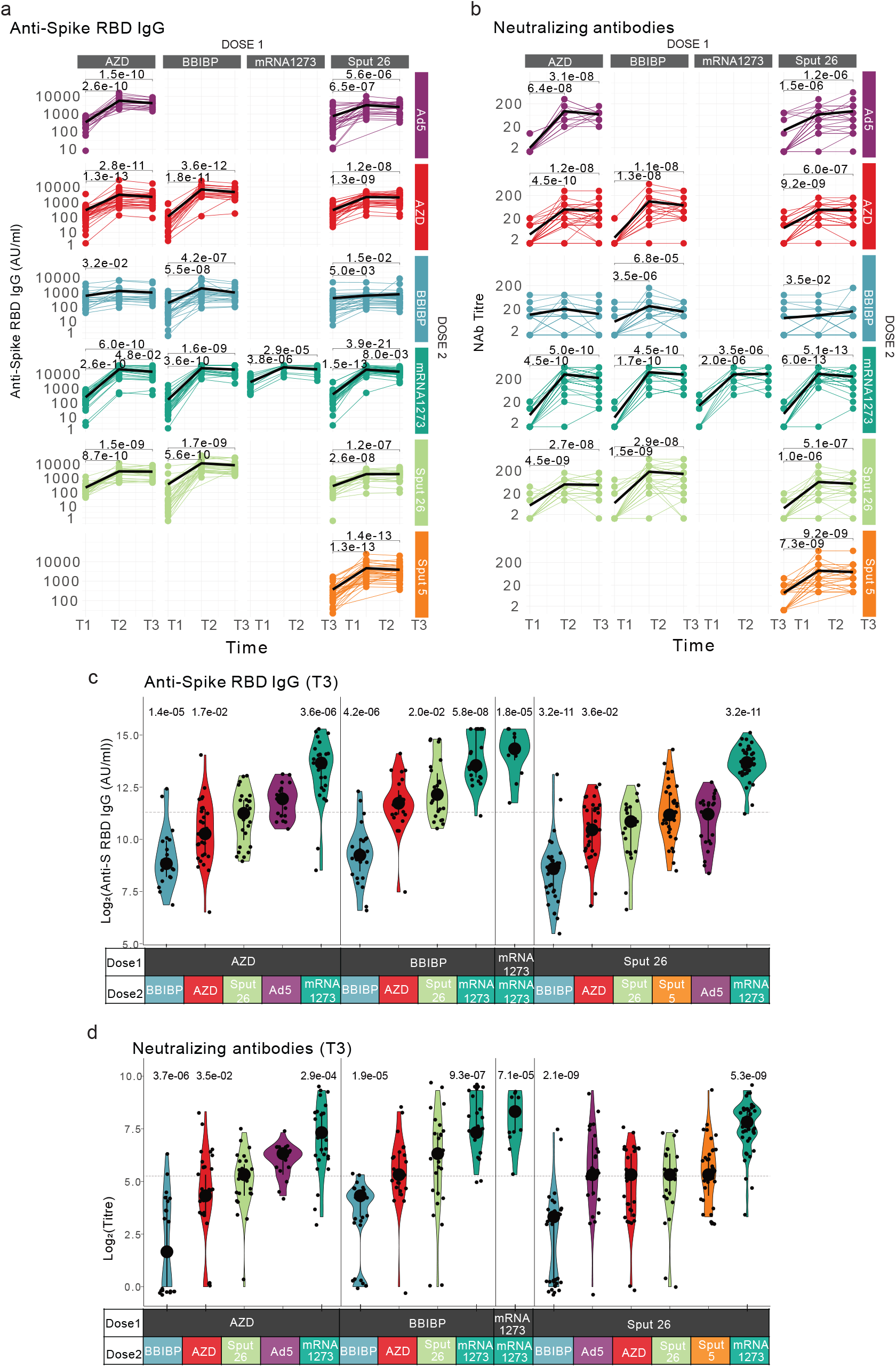
Antibody response in participants after vaccination with various regimens. (a-b) Longitudinal (a) anti-S-RBD IgG levels and (b) NAb titres measured at timepoints T1, T2, and T3. Black lines show the median. (c-d) Log2-transformed (c) anti-S-RBD IgG levels and (d) NAb titres at T3. Large black dots depict the median of each group, and the vertical line spans the interquartile range. The horizontal line shows the overall mean of all participants. P-values (shown above groups) indicate differences between the respective group and the overall mean of all participants. P-values were calculated using the Mann-Whitney-Wilcoxon test and corrected for multiple hypothesis testing with the Benjamini-Hochberg method. Only statistically significant p-values (p < 0.05) are shown.

### Strong B cell response after mRNA-1273

B cells contribute to immunological memory against many viral infections and are important in protecting against COVID-19^35^. The ability of COVID-19 vaccines to induce memory B cell expansion is therefore critical for their efficacy. Hence, we generated a multi-parametric flow cytometry panel to interrogate post-vaccination B cell dynamics, using fluorescently labelled wild-type SARS-CoV-2 spike multimer probes^36^ to identify memory B cells (mBCs) specific for the SARS-CoV-2 spike protein. Data were projected using Uniform Manifold Approximation and Projection (UMAP)^37^ in conjunction with FlowSOM clustering^38^ to evaluate the canonical B cell populations and identify spike-binding mBCs (**Fig. 3a, Extended Data Fig. 3a**). The proportions of all these B cell populations except the spike-binding mBCs were similar among the 16 vaccinated groups at T3 (**Fig. 3b**). One month after Dose 2 (T3), we observed a significant increase in the frequency of this population in all vaccine groups, except for those with BBIBP as Dose 2 (**Fig. 3c**). In line with our findings from the anti-S-RBD IgG and NAb analyses, the most pronounced increase in spike-binding mBCs was observed in individuals who received mRNA-1273 as Dose 2 (**Fig. 3d, Extended Data Fig. 3b**, and **Extended Data Table 1**). Of note, we found that the frequency of these cells correlated with the levels of anti-S-RBD IgG and NAbs for some (but not all) vaccine groups, suggesting that this frequency could serve as an indicator for the systemic antibody response in certain cases, at least at early time points after vaccination (**Fig. 3e**). We did not observe a correlation between the antibody levels and the frequencies of other B cell subsets in most of the vaccine groups (**Extended Data Fig. 3c**).

**Figure 3.**
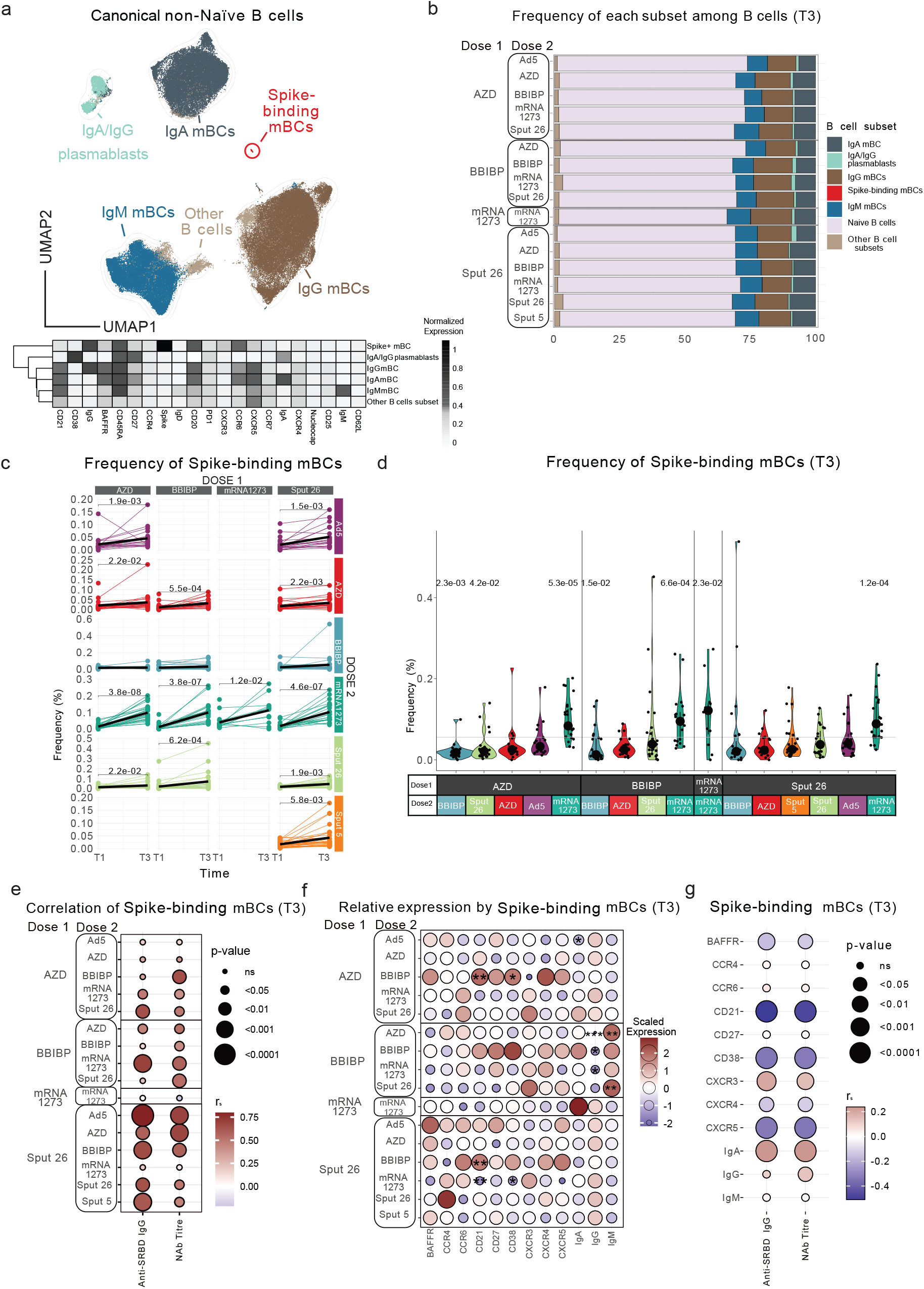
Characterization of spike-binding mBCs from participants receiving different vaccine regimens. (a) UMAP showing the FlowSOM-guided manual metaclustering of non-naïve B cells (IgD^-^/IgM^-^) for all vaccine groups. The heatmap indicates the median intensity of normalized marker expression (range: 0-1) for the identified B cell subsets. (b) Relative frequencies of B cell subsets from each vaccine regimen. (c) Longitudinal analysis of spike-binding mBC frequencies among total B cells between timepoints T1 and T3. Black lines show the median. (d) Frequencies of spike-binding mBCs among total B cells at T3. Large black dots show the median of each group, and the vertical line spans the interquartile range. The horizontal line indicates the overall mean of all participants. P-values (shown above groups) indicate differences between the respective group and the overall mean of all participants. (e) Correlations of spike-binding mBC frequencies with anti-S-RBD IgG levels and NAb titres for each vaccine regimen. (f) The scaled expression of phenotypic markers by spike-binding mBCs at T3 for each group. P-values indicate differences between the respective group and the overall mean of all participants for each marker. *p < 0.05, **p < 0.01, ***p < 0.001. (g) Correlations between phenotypic marker expression by spike-binding mBCs and antibody response (anti-S-RBD IgG levels and NAb titres) across all vaccine regimens at T3. (e,g) Colour indicates the Spearman’s rank correlation coefficient (rs), and the bubble size indicates the p-value. P-values for (b-d,f) were calculated using the Mann-Whitney-Wilcoxon test and corrected for multiplehypothesis testing with the Benjamini-Hochberg method. Only statistically significant p-values (p < 0.05) are displayed.

To better understand the effect of various vaccine regimens on the phenotype of spike-binding mBCs, we evaluated these cells’ expression of differentiation markers and trafficking molecules at T3 (**Fig. 3f**). Spike-binding mBCs from BBIBP-primed groups showed the lowest relative expression of IgG and the highest of IgM, indicating limited class switching (i.e. from IgM to a IgG/IgA isotype). The highest intensity of IgA was found in spike-binding mBCs from individuals vaccinated with mRNA-1273/mRNA-1273 (**Fig. 3f, Extended Data Table 2**). Next, we examined whether these phenotypes were indicative of the strength of the antibody response (**Fig. 3g, Extended Data Fig. 3d**). We found strong negative correlations between antibody response (anti-S-RBD IgG levels and NAb titres) and the expression of CD21, CD38, and CXCR5 on spike-specific mBCs, while IgA and CXCR3 were positively correlated with antibody response. Thus, not only the frequency but also the phenotype of antigen-specific mBCs can reflect the strength of the antibody response after COVID-19 vaccination (**Fig. 3g**). Overall, our results highlight three key points. First, spike-binding mBC frequencies increased after Dose 2 in almost all groups, with the strongest expansion seen in mRNA-1273-boosted individuals. Second, boosting with BBIBP did not significantly increase spike-binding mBC frequencies and resulted in mBCs compatible with a switched resting (CD21^+^CD27^+^CD38^+/lo^) or pre-switched (IgM^+^CD21^+^CD27^+^CD38^+/lo^) phenotype. Boosting with mRNA-1273, meanwhile, induced phenotypes indicating switched activated (IgG/IgA^+^CD21^-^CD27^+^CD38^-^) or atypical (IgG/IgA^+^CD21^-^CD27^-^CD38^lo^) mBCs^39^. Third, spike-binding mBCs from BBIBP-primed individuals showed expression profiles suggesting limited isotype switching (indicated by higher IgM expression) alongside phenotypic markers (CD21, CD38, CXCR5, CXCR3) that correlate with the level of antibody production after vaccination. Thus, the B cell immune landscape in vaccinated individuals clearly mirrors the antibody profile seen in the serum shortly after vaccination.

### BBIBP priming induces highest T cell responses

Increased frequencies of interferon-gamma (IFNγ)-secreting T cells against SARS-CoV-2 spike, nucleoprotein, and matrix proteins are known to predict protection after vaccination from COVID-19^2,35^. To assess the cellular immune response upon antigen re-encounter, we stimulated PBMCs from timepoints T1 and T3 with SARS-CoV-2 spike and nucleocapsid peptide pools and measured IFNγ production with an ELISpot assay (**Fig. 4a-b**). Spike-induced IFNγ production was significantly increased (1.6-fold) at T3 in the AZD/mRNA-1273 group, while BBIBP-primed individuals showed significant increases with all booster vaccines (**Fig. 4a** and **c, Extended Data Fig. 4a**). For Sput 26-primed individuals, only Sput 5 (2.7-fold) as Dose 2 resulted in a significant increase of spike-specific responses at T3. The strongest response in AZD-primed individuals was observed in the AZD/Ad5 group, while BBIBP-primed individuals reached high levels with all vaccine combinations (**Fig. 4c, Extended Data Fig. 4a**, and **Extended Data Table 1**). Meanwhile, the nucleocapsid-induced IFNγ production at T3 in BBIBP-primed participants was highest after boosting with mRNA-1273, BBIBP, or AZD, whereas a Sput 26 boost resulted in a lower response (**Extended Data Fig. 4b**).

**Figure 4.**
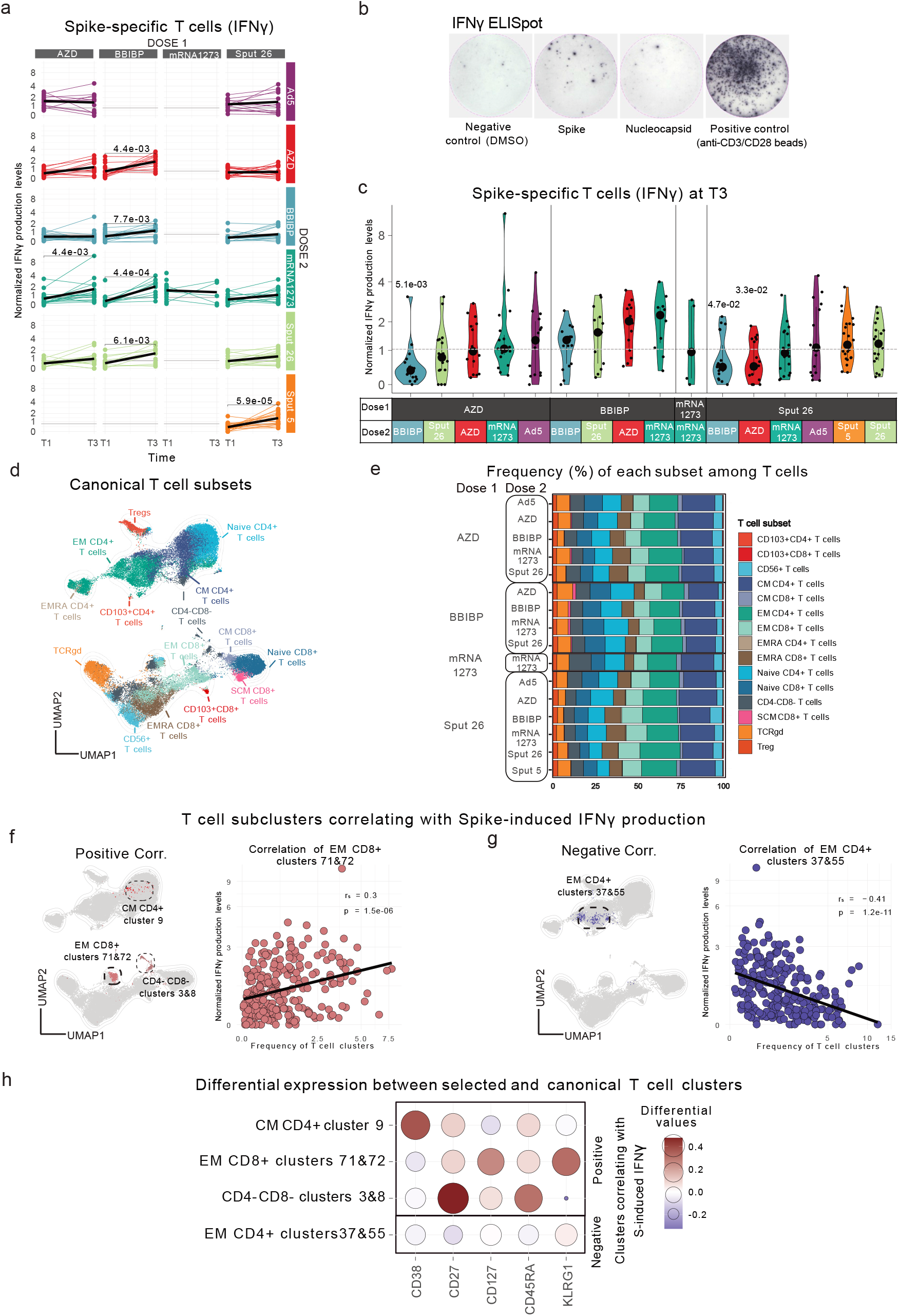
Spike-specific T cell responses to antigen re-encounter after various vaccine regimens. (a,c) Normalized IFNγ production after stimulation of PBMCs with SARS-CoV-2 spike peptide pool, measured by ELISpot assay. (a) Longitudinal IFNγ response at T1 and T3. Black lines show the median. (b) Representative images of one IFNγ ELISpot assay (n=583). (c) Normalized IFNγ responses at T3 after stimulation with SARS-CoV-2 spike peptide pool. Large black dots show the median of each group, and the vertical line spans the interquartile range. The horizontal line indicates the positive cut-off threshold. P-values indicate differences between the respective group and the overall mean of all participants. (a,c) P-values were calculated using the Mann-Whitney-Wilcoxon test and the Benjamini-Hochberg method to control for multiple hypothesis testing. Only statistically significant p-values (p < 0.05) are shown. (d) UMAP showing the FlowSOM-guided manual metaclustering of T cells (CD3+) for all vaccine groups combined. (e) Frequencies of canonical T cell subsets relative to the total number of T cells for each vaccine regimen. (f,g) UMAP of the T cell compartment showing the clusters with frequencies (f) positively or (g) negatively correlated with SARS-CoV-2 spike peptide-induced IFNγ response. (h) Differential marker expression by the specific T cell subclusters compared to the canonical T cell subsets that positively and negatively correlated with SARS-CoV-2 spike peptide-induced IFNγ responses, filtered for markers with at least 0.2 differential expression. The colour shows the Spearman’s rank correlation coefficient (rs), and the bubble size shows the p-value. Only significantly correlated clusters (p < 0.00001) are displayed.

Overall, BBIBP-primed individuals showed the highest spike-induced IFNγ production, independent of Dose 2. BBIBP is the only vaccine in the study that targets the whole SARS-CoV-2 virus rather than solely the spike protein^28^. Therefore, as expected, this was the only vaccine that induced strong cellular immunity against nucleocapsid peptides. Interestingly, we did not detect responses above the threshold level when BBIBP was given as Dose 2 in heterologous regimens (**Fig. 4c**). In sum, these results reveal that T cell-mediated responses against SARS-CoV-2 spike and nucleocapsid peptides are stronger when BBIBP is administered as Dose 1, regardless of the vaccine used as Dose 2.

### T cell clusters track antigen-specific responses

Given the differences in the T cell responses, we interrogated overall T cell dynamics to determine whether the frequencies of specific T cell subsets are indicative of strong spike-specific cellular responses upon antigen re-encounter. We began by generating a lymphocyte-focused panel for single-cell analysis and defined 15 canonical T cell subpopulations using naïve/memory-associated markers (**Fig. 4d, Extended Data Fig. 5a-b**). No significant differences were observed in the relative frequencies of these subsets among the vaccinated groups at T3 (**Fig. 4e**), and none of the subsets showed a strong correlation (|r_s_| > 0.25 and p < 0.05) with Spike-induced IFNγ production (**Extended Data Fig. 5c**). To investigate the canonical T cell subsets more deeply, we reclustered the samples using 27 functional and lineage-specific spectral flow parameters (**Extended Data Fig. 5a**) and performed FlowSOM clustering on the T cell compartment (**Extended Data Table 1**). The frequencies of four of the resulting T cell clusters correlated with the levels of spike-induced IFNγ production detected in the ELISpot assay (|r_s_| > 0.25 and p < 0.05) (**Extended Data Table 3, Fig. 4f-h, Extended Data Fig. 5d**). The phenotypes and the differentially expressed markers of the correlating clusters are depicted in **Extended Data Fig. 5e** and **Fig. 4h**, respectively. CM CD4^+^ cluster 9, EM CD8^+^ clusters 71&72, and CD4^-^/CD8^-^ clusters 3&8 respectively expressed high levels of CD38, KLRG1, and CD27 compared to the canonical T cell populations (**Fig. 4h**). Next, we compared the frequency of these T cell clusters across the 16 vaccine regimens, finding that BBIBP-primed combinations showed higher frequencies of CD4^-^/CD8^-^ clusters 3&8 compared to the other groups (**Extended Data Fig. 5f** and **Extended Data Table 4)**. In sum, we identified post-vaccination T cell clusters that are markers for an antigen-specific T cell response upon antigen re-encounter and are differentially expressed in BBIBP-primed individuals.

**Figure 5.**
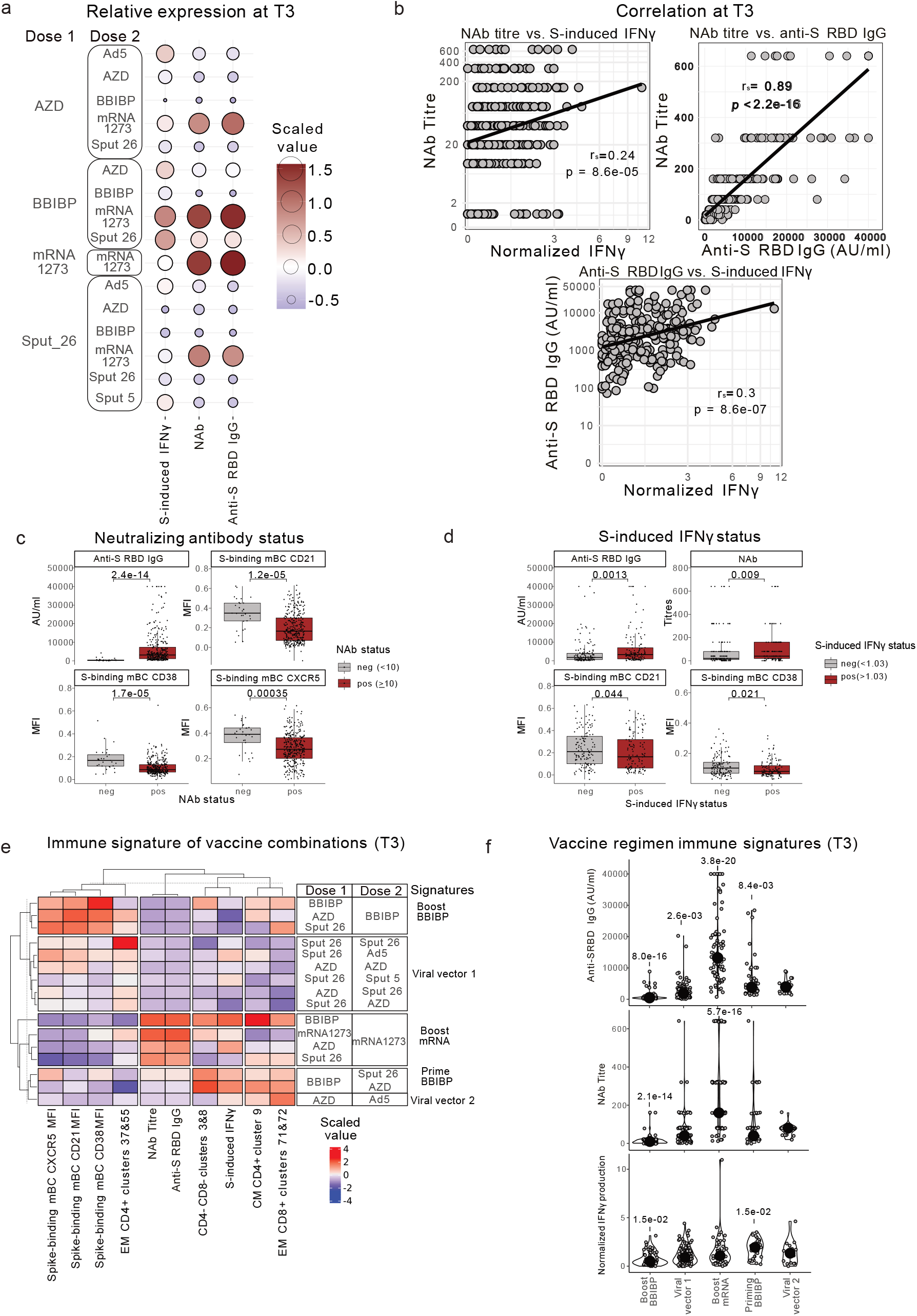
Immune signatures of specific vaccine combinations. (a)Scaled and centred levels of neutralizing (NAb) and spike-specific (anti-S-RBD IgG) antibodies as well as SARS-CoV-2 spike peptide-induced T cell IFNγ production for each vaccine combination at T3. (b) Spearman’s rank correlations between NAb titres, anti-S-RBD antibodies, and SARS-CoV-2 spike peptide-induced IFNγ production. (c-d) Participants were classified as negative (< 10) or positive (≥ 10) for NAb titres (c) and negative (< 1.03, the detection threshold) or positive (> 1.03) for IFNγ response (d), and mean antibody levels and spike-binding mBC marker expression levels were compared between negative and positive participants. MFI = normalized mean fluorescent intensity. Boxes bound the interquartile range (IQR) divided by the median, and Tukey-style whiskers extend to a maximum of 1.5 × IQR beyond the box. Dots are participant data points. (e) Scaled and centred values per column of the top humoral and cellular immune features for each immune signature, displayed in a heatmap with k-means clustering applied to the rows and columns. (f) Anti-S-RBD IgG levels, NAb titres, and SARS-CoV-2 spike peptide-induced IFNγ production from all participants, grouped by immune signature. Large black dots depict the median of the group, and the vertical line spans the IQR. P-values indicate differences between the respective group and the overall mean of all participants. (c-d,f) P-values were calculated using the Mann-Whitney-Wilcoxon test and the Benjamini-Hochberg method to correct for multiple hypothesis testing. Only significant p-values (p < 0.05) are displayed.

### Vaccine regimen immune signatures

Side-by-side analysis of our humoral and cellular immunogenicity data (**Fig. 2c-d, Fig. 4c**, and **Extended Data Table 1**) revealed that BBIBP/mRNA-1273 induced especially high responses in both immune compartments (**Fig. 5a**). We then analyzed whether these primary humoral and T cell parameters correlated with each other. We found a significant correlation between anti-S-RBD IgG levels and spike-induced IFNγ production at T3, while NAb titres were less strongly correlated with the cellular response (**Fig. 5b**). Thus, higher antibody levels (especially anti-S-RBD IgG) are accompanied by higher T cell responses after COVID-19 vaccination, although this correlation was not shared by all vaccine groups (**Extended Data Fig. 6a**).

In our cohort, we observed interindividual differences in the strength of the immune response after vaccination (**Fig. 2c-d** and **Fig. 4c**). Furthermore, the specific immune factors that determine the protective efficacy of COVID-19 vaccines are not entirely understood. In order to assess the immune compositions associated with relatively stronger or weaker immunogenicity, we divided the cohort into positive and negative responders for NAbs and spike-induced IFNγ production (anti-S-RBD IgG status was not included, since all participants were positive responders after Dose 2). Positive responders were defined as neutralizing titre (NAb) ≥ 10 for NAbs and > 1.03 for spike-induced IFNγ production. The proportions of positive responders among the vaccine groups are shown in **Extended Data Fig. 6b-c**. Consistent with **Fig. 3g**, positive responders for NAb titres had higher anti-S-RBD IgG levels and lower expression of CD21, CD38, and CXCR5 on spike-binding mBCs (**Fig. 5c**). Positive responders for spike-induced IFNγ production had significantly higher anti-S-RBD IgG levels and NAb titres and lower expression of CD21 and CD38 by spike-binding mBCs (**Fig. 5d**). In addition, the frequencies of the four T cell clusters that correlated with the IFNγ response (**Fig. 4f-h**) were all significantly different between positive and negative responders for spike-induced IFNγ production (**Extended Data Fig. 6e**) but were similar between positive and negative responders for NAb titres (**Extended Data Fig. 6d**). Thus, higher post-vaccination anti-S-RBD IgG levels and lower expression of CD21 and CD38 by spike-binding mBCs were associated with strong humoral neutralizing activity and antigen-specific T cell responses.

We then combined the mBC response markers from **Fig. 5c-d** and the T cell clusters shown in **Extended Data Fig. 6d-e** in order to better characterize the specific cellular immune profiles associated with the 16 vaccine regimens. K-means clustering resulted in the segregation of five main vaccine regimen signatures (i.e., boost BBIBP, boost mRNA, prime BBIBP, viral vector 1, and viral vector 2) clearly separated through the comparison of 10 parameters (**Fig. 5e**). BBIBP/mRNA-1273, which was the most immunogenic combination with regards to both the antibody and T cell responses, presented its own distinct immune signature characterized by spike-binding mBCs with an activated phenotype (low expression of CD21, CD38, and CXCR5), low frequency of T cells in effector memory (EM) CD4^+^ clusters 37&55, and high frequencies of T cells in CD4^-^/CD8^-^ clusters 3&8, central memory (CM) CD4^+^ cluster 9, and EM CD8^+^ clusters 71&72. These analyses also revealed that while BBIBP was the least immunogenic booster vaccine, it elicited the highest expression of CXCR5, CD21, and CD38 on spike-binding mBCs (all shown to be higher in negative NAb titre responders in **Fig. 5c**). These same markers were all expressed with a lower intensity in individuals boosted with mRNA-1273. Finally, we compared antibody production and cellular immune responses to SARS-CoV-2 among these five groups. The groups “boost mRNA” and “priming BBIBP” respectively induced the highest humoral and antigen-specific T cell responses (**Fig. 5f** and **Extended Data Table 1**). Our analysis thus allows the extraction of vaccine regimen-driven immune signatures, which are linked to immunogenicity. These results provide insights into the underlying immunological mechanisms of vaccine-induced immune responses.

## Discussion

The plethora of different COVID-19 vaccines available offer an unprecedented opportunity to study human immune responses to immunization. Here, we present the most comprehensive head-to-head immunophenotyping comparison of vaccine protocols to date, covering adenoviral-vector, inactivated virus, and mRNA platforms. We show that several heterologous vaccine combinations have similar or superior humoral and cellular immunogenicity compared to homologous regimens. In addition, we classified the 16 vaccine combinations into five distinct groups based on differing humoral and cellular immune signatures induced in vaccinated individuals.

Our finding that the heterologous regimens AZD/mRNA-1273 and AZD/Ad5 induced stronger humoral and cellular responses compared to AZD/AZD agree with preliminary reports showing improved immunogenicity for AZD when combined with mRNA vaccines (mRNA-1273 or BNT162b2)^3,4,7,8,11,12,40^. In BBIBP-primed individuals, all heterologous combinations led to stronger immune responses than the homologous regimen. These findings corroborate the results of antibody analyses from the complete ECEHeVac cohort (which included three additional centres in Argentina)^32^. In general, the administration of mRNA-1273 as Dose 2 clearly improved immunogenicity in all assessed conditions. Combinations such as AZD/Ad5, BBIBP/Sput 26, and BBIBP/AZD also proved to be highly immunogenic. Together, these results provide a strong rationale for the superiority of heterologous vaccine regimens against SARS-CoV-2 among non-mRNA based vaccines, suggesting that this strategy could improve the efficiency of vaccination programmes, particularly in regions with limited vaccine supply. Interestingly, as also observed in other studies, the order of vaccine administration in heterologous regimens also appears to be important – in line with other studies^40–42^, we observed that BBIBP induced strong immune responses as Dose 1 but not as Dose 2.

Spike-binding mBC phenotypes differed among the vaccine combinations; boosting with BBIBP induced a resting phenotype, while boosting with mRNA-1273 led to an activated phenotype^39^. The exact mechanisms underlying these differences in mBC induction remain unclear, but differential interactions between B cells and membrane-bound vs. soluble antigens may be responsible^43^. Furthermore, we found that boosting with BBIBP coincided with the highest proportions of CXCR5^hi^CXCR3^lo^ mBCs, a phenotype reminiscent of naïve B cells. As in our study, Zhang *et al*. reported a correlation between NAb titres and CXCR3 expression by mBCs in individuals who received an adenovirus-based COVID-19 vaccine^44^.

mBCs are of particular interest in the context of new SARS-CoV-2 variants, as these cells undergo fewer somatic hypermutations than plasma cells and are potentially more flexible in responding to different viral subtypes^45^. IgM^+^ mBCs tend to migrate to B cell follicles and re-initiate germinal centre reactions upon rechallenge, thus potentially increasing the breadth of the antibody response; meanwhile, IgG^+^ mBCs preferentially differentiate into plasma cells to rapidly induce specific antibody production^46–48^. In our study, IgM-expression of spike-binding mBCs was higher in BBIBP-primed individuals at T3, independent of the booster. Conversely, priming with the other vaccines resulted in the generation of primarily IgG^+^ spike-binding mBCs. In mouse models, rapamycin-induced reduction of germinal centre formation and loss of class switching in B cells led to broader protective antibody response against influenza virus subtypes^49^, and immunization with Dengue virus variant proteins predominantly stimulated IgM^+^ mBCs^43,50^. In humans, one study showed fewer IgG^+^ cells among switched (IgD^-^) S-RBD-specific mBCs and stronger neutralizing activity against SARS-CoV-2 (including viral variants) after heterologous AZD/BNT162b2 compared to homologous BNT162b2/BNT162b2 vaccination^4^. Thus, a vaccine response that induces less IgG switching of mBCs might lead to a broader immune response, increasing immunity against viral variants. Concordantly, another recent study of homologous vs. heterologous vaccine combinations showed that BBIBP/mRNA-1273 had the highest neutralizing activity against the Omicron variant^32^.

Regarding the induction of spike-specific T cells, priming with BBIBP proved to be an optimal base for a strong IFNγ response. We found two T cell clusters (CD4^-^/CD8^-^ clusters 3&8) that consistently expanded to a greater extent in the BBIBP-primed groups. These clusters were similar to the canonical and CD4^-^/CD8^-^ T cell subsets but displayed higher expression of CD27 and CD45RA and lower expression of KLRG1. While KLRG1^hi^ T cells are generally considered short-lived, it has been shown that KLRG1 can be downregulated (ex-KLRG1), giving rise to multiple memory populations that contribute to an effective antiviral response^51^. Of note, our data do not show whether the described T cell clusters are themselves responsible for the observed IFNγ production after stimulation with spike peptides, as the clusters were identified through correlative analyses. However, they could nonetheless serve as biomarkers for the cellular response after COVID-19 vaccination.

While clinical studies are needed to determine whether the comparatively higher immunogenicity of specific vaccination regimens translates to reliable protection from SARS-CoV-2, the immune signatures described in our study can serve as a guide for vaccine development. It was previously reported that serum antibody titres are not reliable markers of the CD4^+^ T cell response^35^; however, we observed a significant correlation between anti-S-RBD IgG levels and spike-induced IFNγ production 4 weeks after Dose 2. A certain sample size may be needed to detect this relationship, since analysis of individual groups did not reveal such a correlation. The usefulness of IgG antibodies as a marker for the cellular response to COVID-19 vaccines might therefore be restricted to larger cohorts and early time points after vaccination.

A limitation of this study is the difference in time intervals between Doses 1 and 2 for some groups. A 12-week interval was previously shown to induce higher NAb titres, but weaker T cell responses, compared to a 4-week interval for homologous and heterologous combinations of AZD and BNT162b2^52^. Nonetheless, some of the regimens with the shortest dosing intervals in our cohort (e.g., mRNA-1273/mRNA-1273) displayed the highest NAb titres among all the groups, and the intervals were comparable among individuals receiving the same Dose 1. Additionally, we did not evaluate the real-world efficacy of the different vaccine combinations and the persistence of their immune responses over time.

As many of the heterologous prime-boost regimens we investigated had not been previously studied in detail, these results considerably expand our understanding of the most potent combinations of COVID-19 vaccines and provide guidance for the design of clinical trials measuring the effectiveness of these heterologous regimens. In addition, the identified immune signatures of the most successful vaccine protocols may be highly valuable not only for the development and testing of COVID-19 vaccines but also for the future design, selection, and deployment of vaccines against other important diseases as well as cancer.

## Data Availability

All data produced in the present study are available upon reasonable request to the authors
All data produced in the present work are contained in the manuscript
All data produced are available online at

## Methods

### Sample donors

Volunteers (age range 18-82 years old) were enrolled in a randomized, open Phase IIB clinical trial (ECEHeVac, NCT04988048) aimed at comparing the immunogenicity and reactogenicity of heterologous and homologous vaccination regimens available in Córdoba, Argentina. The study received ethical approval by the Registro Provincial de Investigación en Salud (Provincial Registry of Health Research, REPIS-Cba #4371). The study was conducted in accordance with the guidelines of Good Clinical Practice (ICH 1996) and the principles of the Declaration of Helsinki.

### Eligibility criteria

Eligible participants were healthy volunteers older than 18 years who had received a first dose of the AZD, BBIBP, Sput 26, or mRNA-1273 vaccine 30-120 days prior to the enrolment date. Exclusion criteria were: immunocompromised status with underlying disease or immunosuppressive treatment; pregnancy and lactation; having received a major surgical intervention in the 30 days prior to the enrolment date; having had a severe allergic reaction (anaphylaxis) to any vaccine; having a visceral disease that lead to disability (heart failure, kidney failure, respiratory failure, liver failure, intestinal malformations, electro-dependence, or having had a visceral transplant less than 2 years previously); and having had COVID-19 (symptomatic or asymptomatic) or a positive anti-nucleocapsid IgG via ELISA on T1 (except for those subjects that had been vaccinated with BBIBP as the first dose).

### Randomization, consent, and follow-up

Participants were randomized (1:1) to determine the vaccine used as Dose 2, and the participant and healthcare personnel in charge of vaccination were informed of the result. Each participant provided written consent to be included in the study. All participants filled out a questionnaire to verify personal data and health history. In this paper, the day of Dose 2 is referred to as time point 1 (T1). The participants were observed for 15-20 minutes after inoculation. After T1, telephone and face-to-face monitoring of each participant was carried out for up to 6 months. The data obtained were recorded in the national health information system (www.https://sisa.msal.gov.ar/sisa/#sisa).

### Serum, plasma, and PBMC collection

Whole blood samples were collected from participants immediately before administration of Dose 2 (T1) and at 14 ± 2 (T2) and 28 ± 1 (T3) days after Dose 2. Sera and plasma were obtained from the whole blood at each timepoint. PBMCs were isolated from samples at T1 and T3 via density-gradient sedimentation using Ficoll-Paque™ PLUS (GE Healthcare). Isolated PBMCs were cryopreserved in heat-inactivated foetal bovine serum (FBS; Natocor) containing 10% DMSO (Sigma-Aldrich) and stored in liquid nitrogen until use. Plasma samples were used for SARS-CoV-2 neutralization assays, sera were used for IgG anti-SARS-CoV-2 assays, and PBMC samples were used for flow cytometry and functional T cell assays.

### Detection of anti-SARS-CoV-2 IgG levels

IgG antibodies against the SARS-CoV-2 nucleocapsid protein were qualitatively detected on T1 using a chemiluminescent microparticle immunoassay (CMIA; ARCHITECT SARS-CoV-2 IgG, 6R86, Abbott) that relies on an assay-specific calibrator to report a ratio of specimen to calibrator absorbance (S/C). The interpretation of the result is determined by an index value, which is a ratio over the threshold value. An index (S/C) of < 1.4 was considered negative and ≥ 1.4 was considered positive. IgG antibodies against the receptor-binding domain (RBD) of the S1 subunit of the SARS-CoV-2 spike protein were evaluated in sera samples collected on T1, T2, and T3 by a quantitative CMIA assay (AdviseDx SARS-CoV-2 IgG II, 6S60, Abbott). Sera with values ≥ 50.0 Arbitrary units/ml were considered positive.

### Detection of neutralizing antibodies against live SARS-CoV-2

Plasma samples from T1, T2, and T3 were tested for their ability to neutralize wild-type SARS-CoV-2 B.1 (hCoV-19/Argentina/PAIS-G0001/2020, GISAID accession ID: EPI_ISL_499083) using the plaque reduction neutralization test (PRNT) as previously described^53^. Briefly, this test was performed with Vero 76 cells (ATCC CRL-1587) that were seeded in 24-well plates 48 hours before infection. Plasma samples were heat-inactivated by incubation at 56°C for 20 minutes and centrifuged at 10,000 rpm 30 minutes before use. Treated samples were diluted two-fold, and an equal volume of virus stock containing 100 plaque-forming units (PFU) was added to each corresponding well until reaching final dilutions ranging from 1:10 to 1:320. Cells were incubated with 0.5% agarose with DMEM supplemented with 2% FBS for 4 days at 37°C in a 5% CO2 incubator. After 4 days, cells were fixed and inactivated using a 10% formaldehyde/PBS solution and stained with 1% crystal violet. NAb titres corresponded to the maximum dilution of plasma that neutralized 80% of the PFU, compared with PFU from the viral controls included in the test.

### IFNγ ELISpot assay

IFNγ ELISpot analysis was performed using microplates pre-coated with monoclonal IFNγ-specific antibodies (Human Interferon Gamma ELISPOT Kit, Abcam). PBMCs (250,000 cells/well) were separately stimulated for 18 hours with peptide pools (PepMix SARS-CoV-2 (Spike Glycoprotein) and (NCAP), JPT Peptide Technologies) at a concentration of 1 µg/ml. Tests were conducted with a negative (DMSO, Sigma-Aldrich) and a positive control (Dynabeads Human T-Activator CD3/CD28, Gibco) for each sample. According to the manufacturer’s protocol, we added a biotinylated anti-IFNγ detection antibody, followed by a streptavidin-AP conjugate and a 5-bromo-4-chloro-3-indolyl phosphate (BCIP)/nitro blue tetrazolium (NBT) substrate (all from the Human Interferon Gamma ELISPOT Kit) to visualize bound IFNγ.

Plates were scanned using an AID Classic ELISpot Reader, and spots were counted with the AID ELISpot software version 7.0 (AID Autoimmun Diagnostika GmbH), following guidelines for the automated ELISpot evaluation^54^. Samples were excluded if the negative control wells had more than 39 or the positive control wells fewer than 40 spots. Spot counts were multiplied by 4 to evaluate spots per million cells and normalized by dividing by the well saturation of the positive control (spots per million cells/positive control well saturation) for each sample. We used repetitive control samples for both acquisition rounds to control for batch effects. The positive cut-off threshold was calculated by taking the mean of the normalized IFNγ responses of the negative control (DMSO) across all groups.

### Flow cytometry and data acquisition

Biotinylated full-length spike and nucleocapsid proteins (R&D Systems) were multimerized with streptavidin (SA)-BV421 (200 ng spike with 20 ng SA; ∼4:1 molar ratio) and SA-PE-Cy5 (50 ng nucleocapsid with 14 ng SA; ∼4:1 molar ratio) respectively, for 1 hour^36,55^. For the spike-binding mBC and T cell panels, 1.5×10^6^ and 1.0×10^6^ PBMCs respectively were washed with PBS and blocked using Human TruStain FcX and True-Stain Monocyte Blocker (BioLegend). First, for the spike-binding mBC panel, cells were incubated for 30 minutes at 37°C with the tetramers and antibodies. For both the spike-binding mBC and T cell panels, cells were stained for 25 minutes at 4°C. Following surface staining, cells were fixed with 2% PFA or with Foxp3/Transcription Factor Fixation/Permeabilization solution (eBioscience) for 15 or 40 minutes at 4°C, respectively, for the spike-binding mBC and T cell panels. For the T cell panel, cells were then stained overnight at 4°C, all diluted in 1X Permeabilization Buffer (eBioscience). Data were acquired with Cytek Aurora flow cytometers and preprocessed using FlowJo software version 10 (BD Bioscience).

### High-dimensional flow cytometry data analysis

For high-dimensional flow cytometry analysis, dead cells, doublets, or cells stained by fluorochrome aggregates were excluded from the analysis via manual gating using FlowJo. Datasets of different batches were corrected using the CytoNorm R package^56^. To obtain an unbiased overview, we systematically reduced cytometry data to two dimensions by applying UMAP (umap R package^57^) to stochastically selected cells. All cells were clustered using the FlowSOM algorithm (FlowSOM R package^38^) in conjunction with consensus clustering (ConsensusClusterPlus R package^58^) and were subsequently manually annotated into different clusters with distinct phenotypes in terms of median fluorescence intensity (MFI) of the selected surface marker. The expression of each marker across all samples was min-max normalized to the range 0 to 1. The main R script was run as described by Brummelman *et al*.^59^.

### Statistical analysis

Comparisons of continuous variable means between groups or to the overall mean of all participants were performed using the Wilcoxon-Mann-Whitney test and corrected for multiple hypothesis testing with the Benjamini-Hochberg method (ggpubr R package^60^); these tests were two-tailed and performed on unpaired data. To calculate log-transformed data, we added 1 to the value before taking the log2. Where the y-axis was scaled, the pseudo_log_trans(base = 2) function was used (scales R package^61^). Spearman’s rank correlation coefficients (rs) between continuous variables were calculated with the Hmisc R package^62^, which approximates p-values by using asymptotic t distributions. Scaled expression plots were scaled (values were divided by the standard deviation of the group) and centred (the group mean was subtracted from the values) per marker or feature. Differential expression was calculated by subtracting the expression level of the canonical T cell subset from the expression level of the T cell subcluster of interest. Fold change was calculated by dividing the participant value at T3 by the mean group value at T1. K-means clustering was performed on the rows and columns of heatmaps (ComplexHeatmap R package^63^). P < 0.05 was considered statistically significant. All statistical analyses were performed using R version 4.0.1 (R Core Team 2020).

### Materials availability

This study did not generate new unique reagents.

## Acknowledgements

We thank the study participants who contributed to this work. We thank the authorities of Facultad de Ciencias Químcas-UNC and Dr. Jorge Zarzur (Fundación para el Progreso de la Medicina) for their support to the project. We thank Nicolas Ponce, Paula Alejandra Icely, Gabriela Furlan, and Noelia Maldonado for their technical assistance. This project has received funding from the European Research Council (ERC) under the European Union’s Horizon 2020 research and innovation programme grant agreement No 882424, the Swiss National Science Foundation (733 310030_170320, 310030_188450, and CRSII5_183478 to B.B.), Sinergia 183478 and The Loop Zurich – Medical Research Center, Agencia Nacional de Promoción Científica y Técnica (PICT 2021 CAI-I #00051) and Fondo para la Investigación Científica y Tecnológica (FONCyT) [PICT IP COVID 19-464, 2020]; School of Medical Sciences, National University of Córdoba, Argentina and the Ministry of Health of Córdoba Province, Argentina. N.G.N. is a recipient of a University Research Priority Program (URPP) postdoctoral fellowship. S.K. and T.W. are recipients of a postdoctoral research fellowship of the German Research Foundation (DFG). We thank Dr. Daniel Ackerman from Insight Editing London for critical review and editing of the manuscript.

## Author contributions statement

M.E.P., J.M.C., and C.V. designed the clinical trial (ECEHeVac, NCT04988048). D.C. and G.B. led the clinical trial in Córdoba (REPIS-Cba #4371) and provided funding for the generation of the PBMC biobank. L.L. and P.C. were in charge of the randomization, participant data collection and follow up of donors. C.M, L.O., A.G., C.S., and the ImmunoCovidCba group collected and processed PBMC samples. E.V.A.R., L.C., B.M., G.M., and M.M. supervised the generation of the PBMC biobank. G.C., E.R., S.B., B.K., and the InViV working group performed antibody and neutralizing titre measurements. N.G.N., J.S., and C.A. conceptualised and designed flow cytometry and ELISpot experiments. N.G.N., J.S., C.A., S. Kreutmair, S. Krishnarajah, S.U., J.C.K., S.C.P., F.I., M.S., J.V.V., C.H.M., A.S., T.W., M.L., and D.V. performed experiments. L.P., N.G.N., and J.S. analyzed the data. J.S., N.G.N., L.P., and S. Krishnarajah wrote the original draft. J.S., N.G.N., L.P., S. Krishnarajah, B.B., C.M., G.M., L.C., E.V.A.R., B.A.M., M.M., S. Kreutmair, S.U., F.I., and T.W. reviewed and edited the manuscript. B.B., S.G., G.M., L.C., E.V.A.R., B.A.M., and M.M. supervised and funded the study. All authors read and approved the manuscript.

