## Supplementary figure 1-6 for "Distinct immune signatures discriminate SARS-CoV-2 vaccine combinations"

Extended data Fig. 1

### After the first dose

a

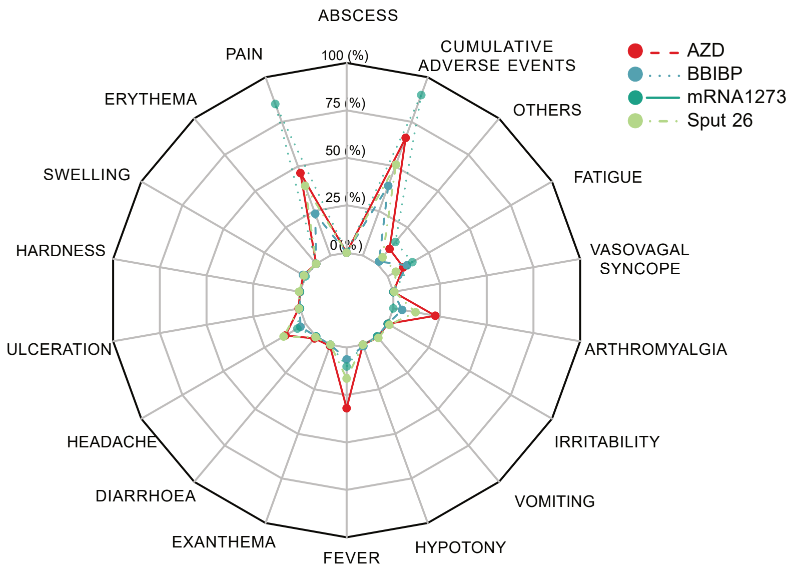


### After the second dose

b

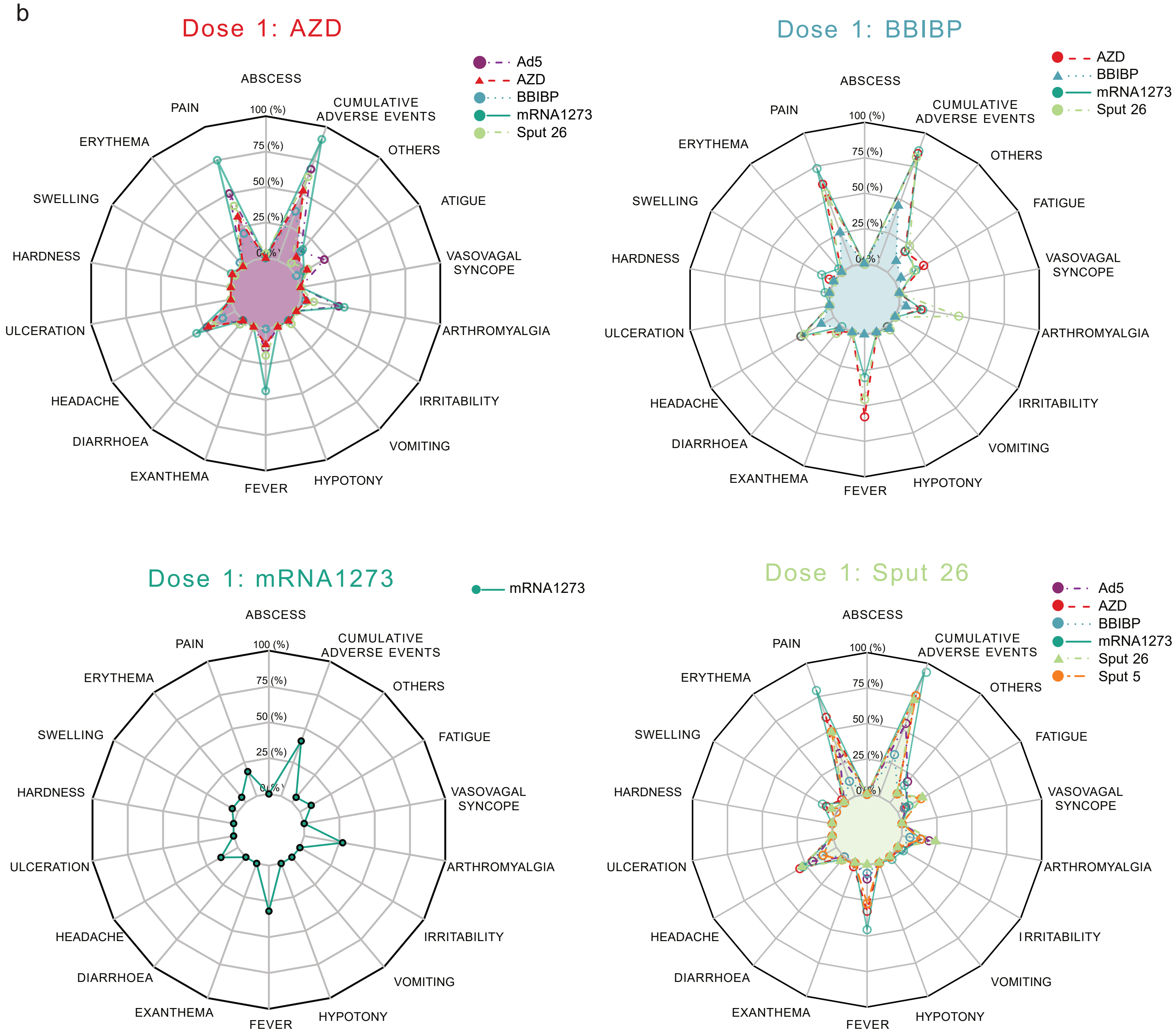

Extended data Fig. 2

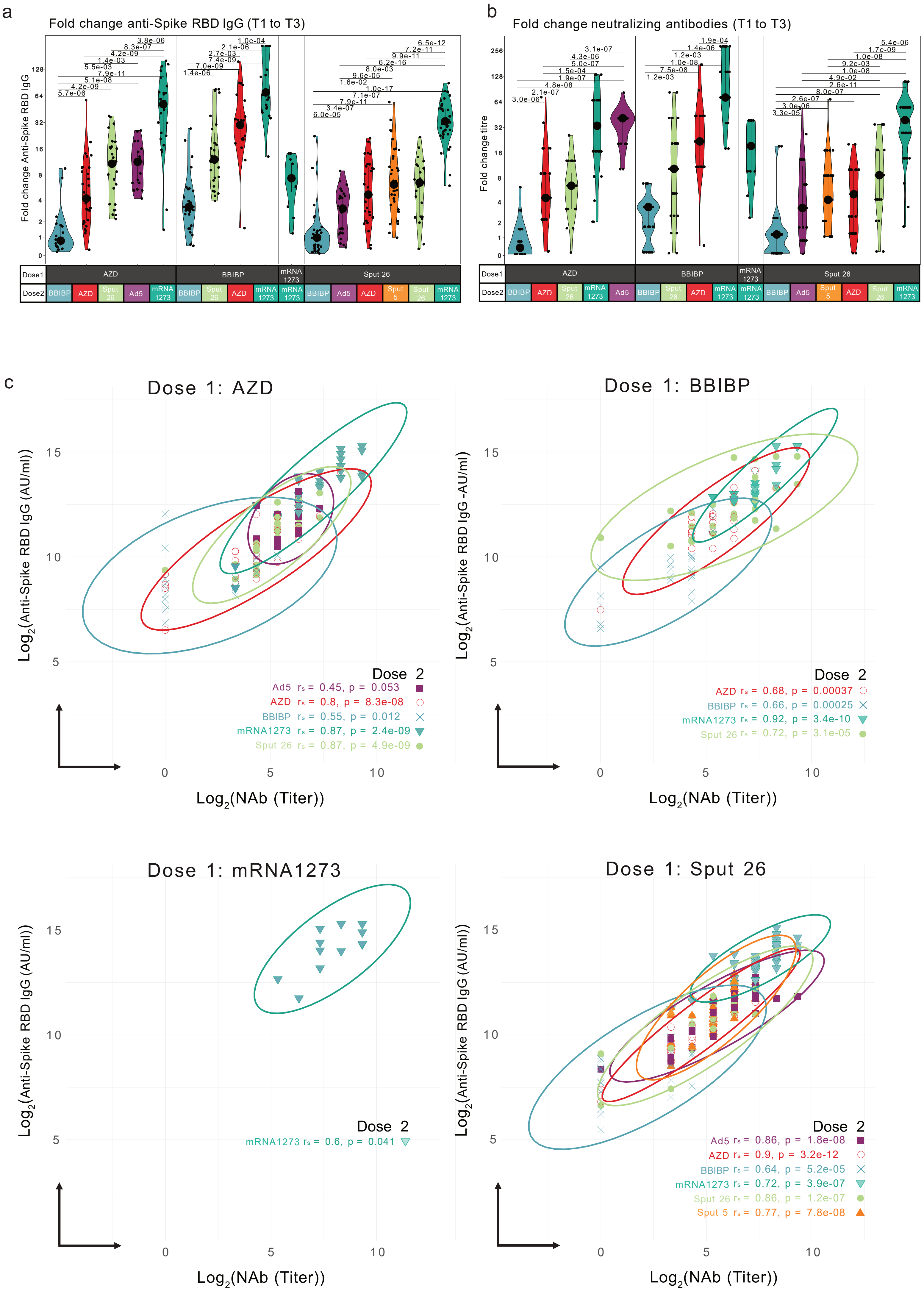

Extended data Fig. 3

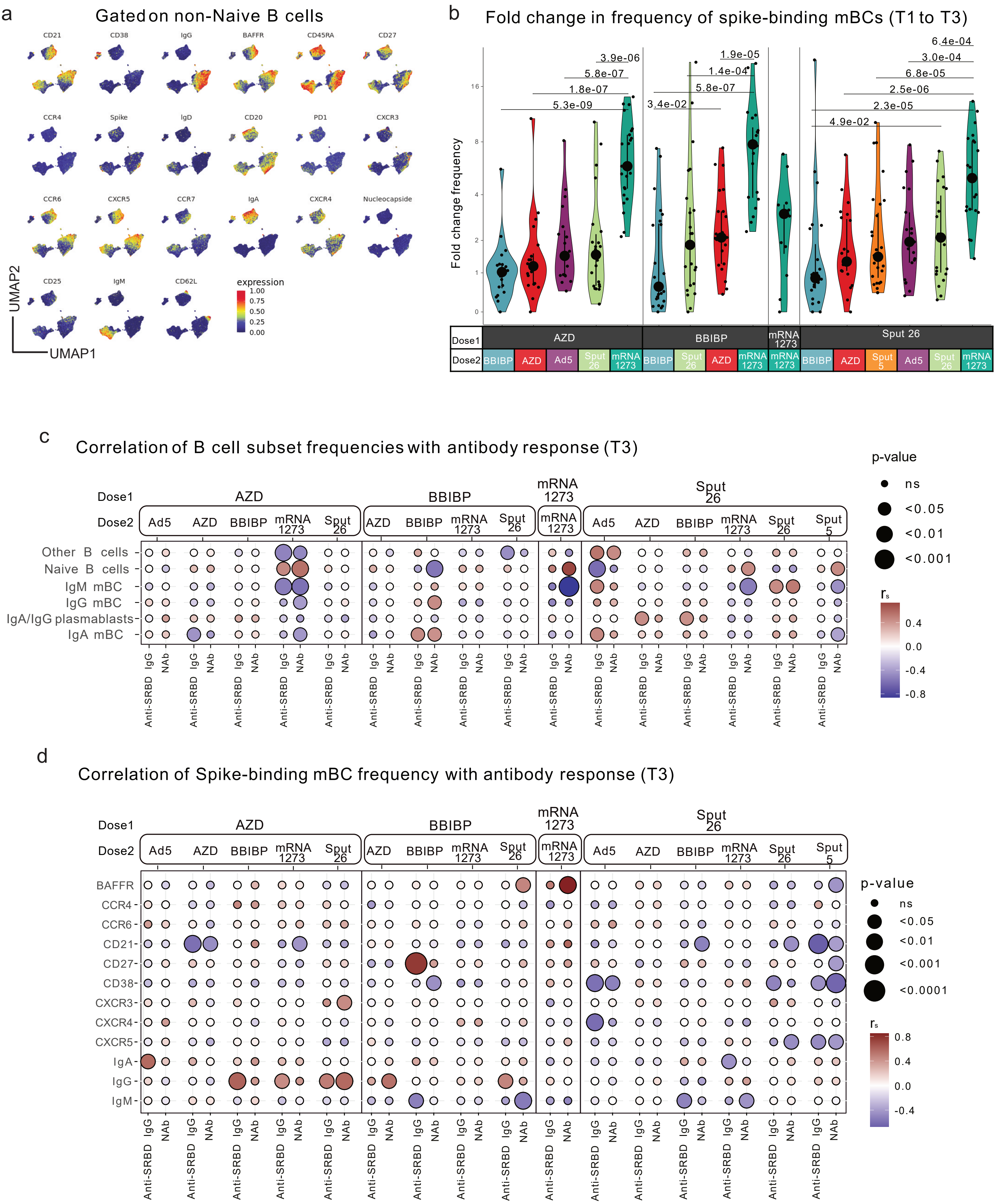

a

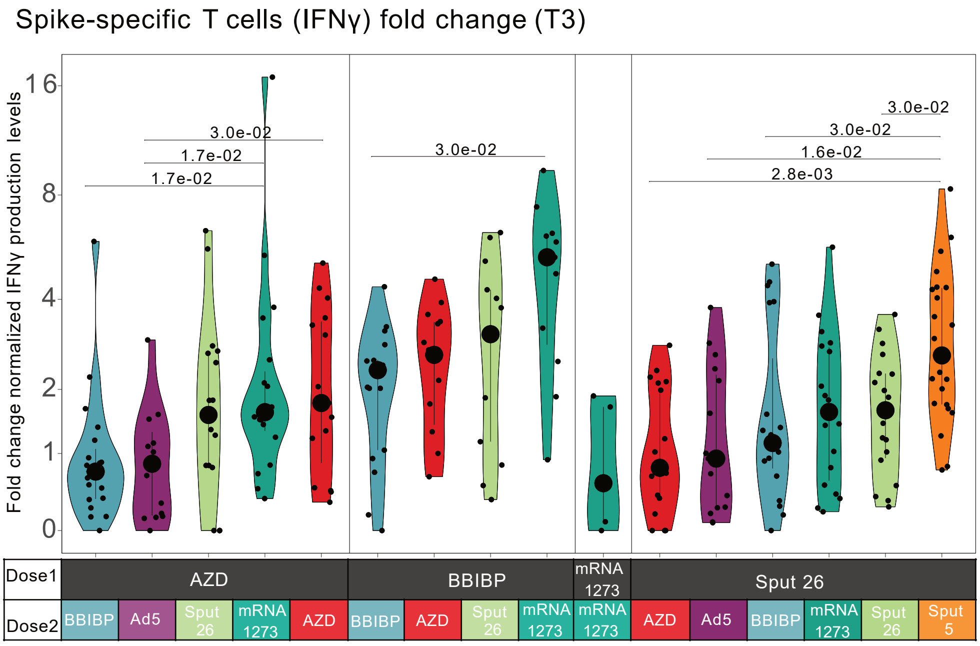
Spike-specific T cells (IFNγ) fold change (T3)
Fold change normalized IFNγ production levels
16
8
4
2
1
0
3.0e-02
1.7e-02
1.7e-02
3.0e-02
3.0e-02
3.0e-02
1.6e-02
2.8e-03
Dose1
AZD
BBIBP
mRNA 1273
Sput 26
Dose2
BBIBP
Ad5
Sput 26
mRNA 1273
AZD
BBIBP
AZD
Sput 26
mRNA 1273
mRNA 1273
AZD
Ad5
BBIBP
mRNA 1273
Sput 26
Sput 5

b

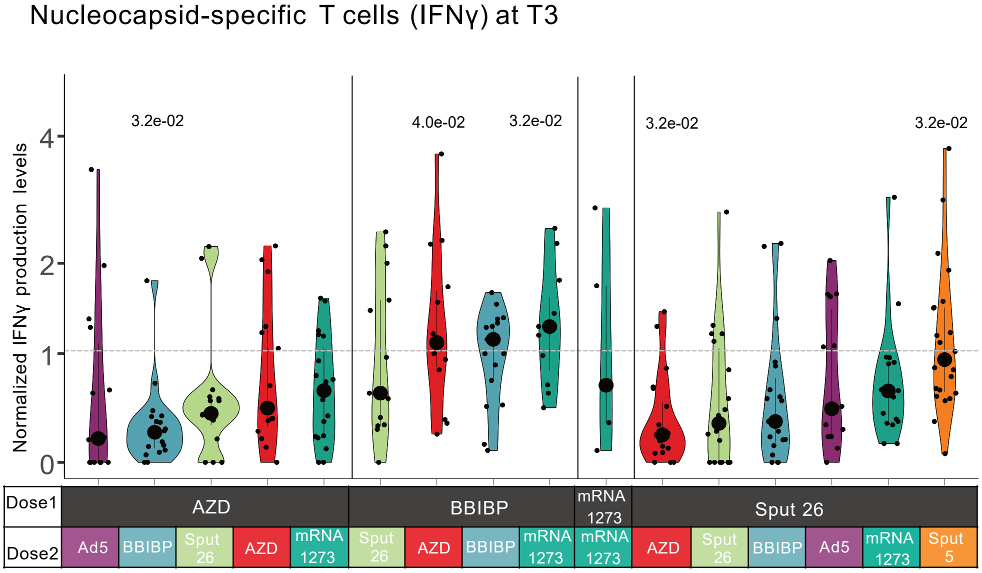
Nucleocapsid-specific T cells (IFNγ) at T3
Normalized IFNγ production levels
4
2
1
0
3.2e-02
4.0e-02
3.2e-02
3.2e-02
3.2e-02
Dose1
AZD
BBIBP
mRNA 1273
Sput 26
Dose2
Ad5
BBIBP
Sput 26
AZD
mRNA 1273
Sput 26
AZD
BBIBP
mRNA 1273
mRNA 1273
AZD
Sput 26
BBIBP
Ad5
mRNA 1273
Sput 5

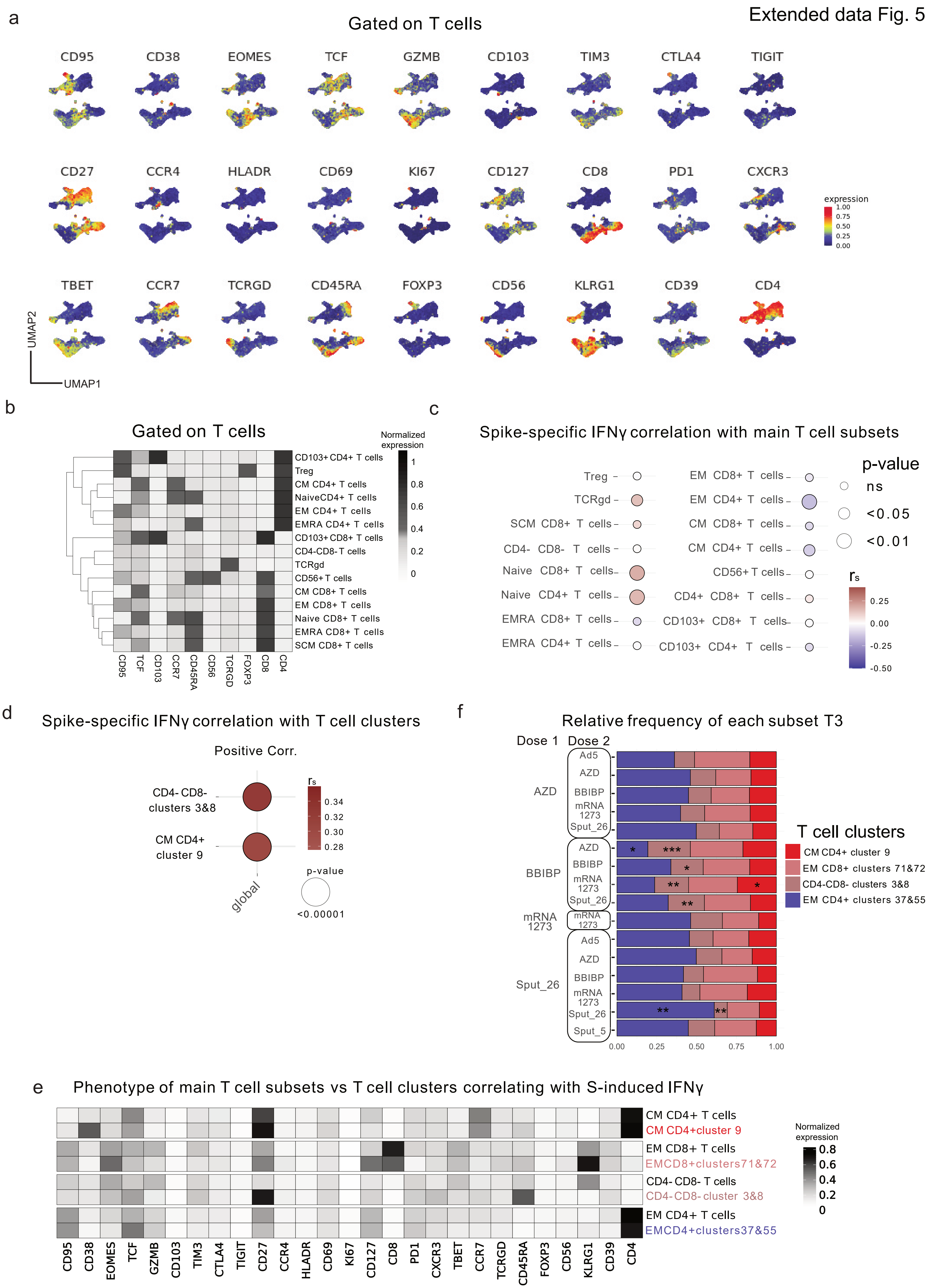

Extended data Fig. 5
a
Gated on T cells
CD95 CD38 EOMES TCF GZMB CD103 TIM3 CTLA4 TIGIT
CD27 CCR4 HLADR CD69 KI67 CD127 CD8 PD1 CXCR3
TBET CCR7 TCRGD CD45RA FOXP3 CD56 KLRG1 CD39 CD4
expression
UMAP2
UMAP1
b
Gated on T cells
Normalized expression
CD103+CD4+ T cells
Treg
CM CD4+ T cells
NaiveCD4+ T cells
EM CD4+ T cells
EMRA CD4+ T cells
CD103+CD8+ T cells
CD4-CD8- T cells
TCRgd
CD56+ T cells
CM CD8+ T cells
EM CD8+ T cells
Naive CD8+ T cells
EMRA CD8+ T cells
SCM CD8+ T cells
c
Spike-specific IFNγ correlation with main T cell subsets
p-value
ns
<0.05
<0.01
d
Spike-specific IFNγ correlation with T cell clusters
Positive Corr.
CD4- CD8- clusters 3&8
CM CD4+ cluster 9
global
f
Relative frequency of each subset T3
Dose 1
Dose 2
T cell clusters
CM CD4+ cluster 9
EM CD8+ clusters 71&72
CD4-CD8- clusters 3&8
EM CD4+ clusters 37&55
e
Phenotype of main T cell subsets vs T cell clusters correlating with S-induced IFNγ
CM CD4+ T cells
CM CD4+cluster 9
EM CD8+ T cells
EMCD8+clusters71&72
CD4- CD8- T cells
CD4-CD8- cluster 3&8
EM CD4+ T cells
EMCD4+clusters37&55
Normalized expression

Extended data Fig. 6

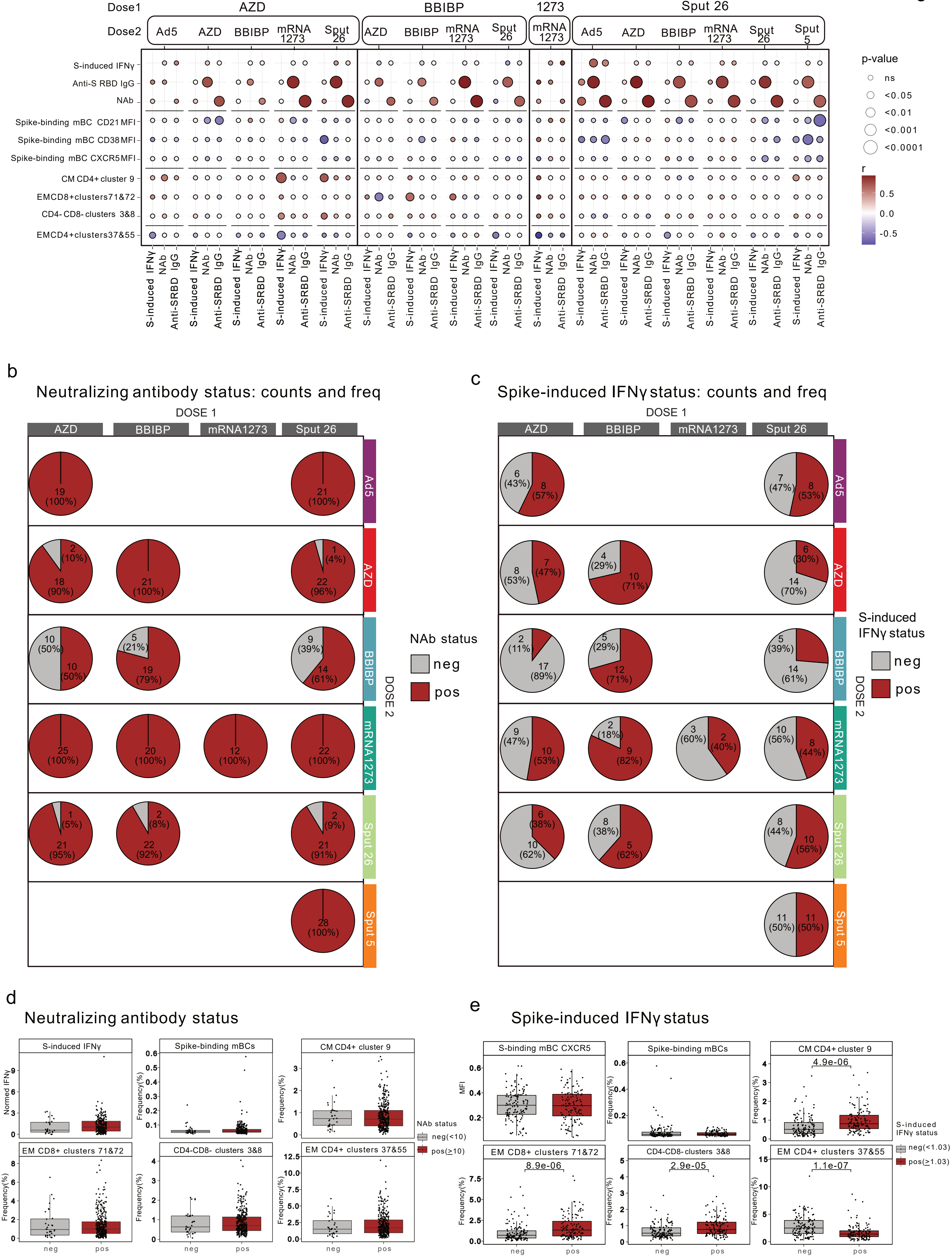
